# redBERT: A Topic Discovery and Deep Sentiment Classification Model on COVID-19 Online Discussions Using BERT NLP Model

**DOI:** 10.1101/2021.03.02.21252747

**Authors:** Chaitanya Pandey

**Affiliations:** Bhartiya Vidyapeeth College of Engineering, GGSIPU, New Delhi, India

**Keywords:** COVID-19, Natural Language Processing, Topic modelling, Deep learning

## Abstract

A Natural Language Processing (NLP) method was used to uncover various issues and sentiments surrounding COVID-19 from social media and get a deeper understanding of fluctuating public opinion in situations of wide-scale panic to guide improved decision making with the help of a sentiment analyser created for the automated extraction of COVID-19 related discussions based on topic modelling. Moreover, the BERT model was used for the sentiment classification of COVID-19 Reddit comments. These findings shed light on the importance of studying trends and using computational techniques to assess human psyche in times of distress.

## 1 Introduction

The internet serves as the most easily accessible source to gauge someone’s mental state and one of the most valuable public expression outlets. Online forums, like Reddit, can be accessed by healthcare service providers, who use users experience data for behaviour analysis and knowledge discovery. Reddit’s interface offers an intuitive way of starting a discussion with other members by joining or creating a new topic, post, or question. By analysing people’s positive and negative comments, or problems and needs related to health issues, we can identify valuable recommendations for improving health services and understanding how to tackle dissent. In late December 2019, the World Health Organization declared a state of emergency due to the rapid spread of the novel coronavirus, causing COVID-19[1].

This paper analyses COVID-19 related comments to detect sentiment and semantic ideas related to COVID-19 based on public opinion with topic modelling. Specifically, machine-learning and NLP were used to derive public sentiments and study the public mindset’s shift throughout a pandemic. This paper’s main contributions are web scraping and topic modelling to extract meaningful topics from COVID-19 related Reddit comments and an in-depth comparison of their polarity, followed by using a deep learning model based on BERT[2] for sentiment classification of COVID-19 related comments. This paper’s findings shed light on the importance of using public opinions and suitable computational techniques to understand issues surrounding COVID-19 and guide related decision-making. Overall, the paper is structured as follows. Section 2 offers a look at some of the similar works. In section 3, the research methodology and pre-processing data methods adopted are discussed, followed by an in-depth presentation of the results obtained on applying deep learning methods to the comment and heading database in section 4. Next, the results are analysed, and we conclude with a discussion on future endeavours based on the proposed model.

## 2. Related Work

The use of NLP for sentiment and semantic analysis to extract meaningful opinions from Twitter, Reddit, and other online health forums has been implemented by many researchers. Martin Müller[3] and colleagues created a transformer model based on BERT, which was trained on a Twitter message corpus. Their model showed a 10-30% improvement over the base model and can be used in several classification and question answering tasks. Sohini Sengupta[4] and colleagues analysed topics related to mental health being discussed on Twitter and used their findings to understand the impact of COVID-19 on mental health amidst the pandemic. Hamed Jelodar[5] and colleagues used automated extraction of COVID-19 related discussions from social media and an NLP method based on topic modelling to uncover various issues related to COVID-19 from public opinions using the LSTM neural network. Halder[6] and colleagues focused on analysing the emotional status of a user over time by studying linguistic changes with the help of a recurrent neural network (RNN) to investigate user-content in a large dataset from online mental health forums. McRoy[7] and colleagues investigated ways to automate the identification of breast cancer survivors’ information needs basing on online health forums. Chakravorti[8] and colleagues evaluated user posts over several subreddits from 2012 to 2018 to extract health issue related topics discussed in online forums. VanDam[9] and colleagues adopted a classification approach for identifying clinic-related posts in online health communities. To create the dataset, they collected 9576 thread-initiating posts from WebMD, a health information website.

Although there are similar works regarding health issues in online forums, to my knowledge, this is the first study to incorporate the BERT model to analyse COVID-19-related comments and headlines from subreddit forums and using topic modelling algorithms to automatically extract meaningful topics and implement a deep learning sentiment classification model to understand the positive and negative opinions related to COVID-19 issues to inform relevant decision-making.

**Fig 1:**
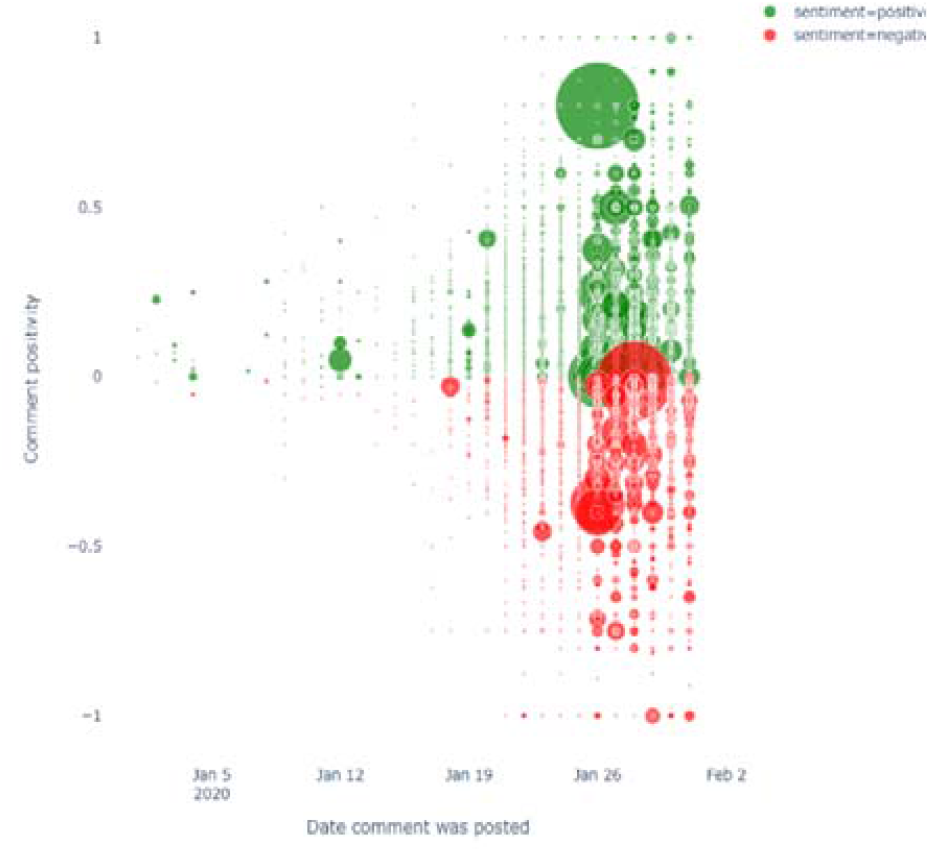
Polarity distribution of Reddit comments.

## 3. Research Methodology

This section breaks down the methods used to achieve this study’s main contributions, proposing a topic model based on unsupervised learning with a collaborative deep-learning model that draws on BERT to analyse COVID-19 related comments in various subreddits.

### 3.1 Obtaining and Preparing the data

#### 3.1.a. Data Collection

Reddit, often coined as “the front page to the internet”, is an American social media website consisting of an extensive network of communities where users can express their opinions via posting questions, comments and communicating to each other regarding various subjects. The posts are organised by online users’ subjects, called “subreddits”, which cover a variety of topics. This distinct categorisation makes Reddit an ideal source for collecting information regarding COVID-19. The unfiltered look at public sentiment makes it the ideal source for analysis. The Reddit API[10] was used to scrape the comments and headlines to add to the dataset, stored in a CSV file updated continuously with checkpoints. This paper focuses on the use of 10 subreddits to create an automated dataset.

#### 3.1.b. Data Pre-processing

One of the most critical pre-processing steps is removing stop words, which are nugatory words that increase dimensionality and space without effectively influencing the output, such as articles, conjunctions, pronouns, and linking verbs. Are, they, these, are some common examples. This is followed by removing special characters, converting words to lowercase, stemming, and lemmatisation, which involves removing suffix and prefixes and converting words to their root form, respectively. Algorithm-1 represents the basic structure of pre-processing input data.

##### Algorithm 1: Input data preparation

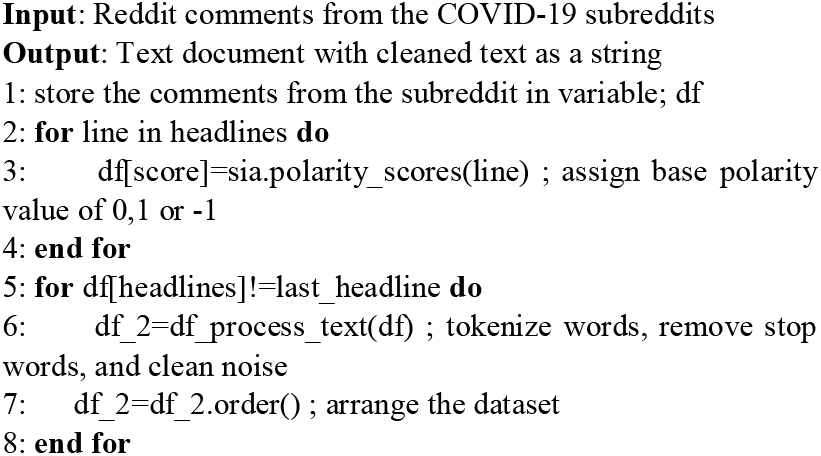

### 3.2. Topic Modelling

Topic modelling is a process that automatically identifies topics present in a text document and derives hidden patterns exhibited by a text corpus. It incorporates an algorithm to extract the key topics in a body of text and build semantic relationships between topics. Topic modelling techniques like Latent Dirichlet Allocation (LDA)[11] and Gibbs Sampling[12] were used for semantic extraction and latent topic discovery of COVID-19 related comments, followed by searching for keywords related to mental health and creating a bigram and network diagram of relevant words to form a general opinion on various issues. The process was visualised with the help of word clouds and graphs.

Regarding the LDA model, the COVID-19 related comments and words are considered topics (K), where the discrete topic distributions are taken from a symmetric Dirichlet distribution. The probability of the observed data (P) was computed and obtained from every COVID-19 related comment in a corpus using the following equation (1):

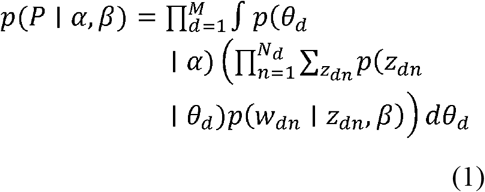

Determined (α) parameters of topic Dirichlet prior and considered word Dirichlet parameters prior (β). M represents the number of text-documents, and N is the length of the vocabulary. Moreover, a pair of Dirichlet multinomials[13] was used to determine (α, *θ*) for the corpus-level topic distributions. Similarly, (β, *γ*) was also determined for the topic-word distributions with a pair of Dirichlet multinomials. Also, the document-level variables are defined as (*θ*_*d*_), which is sampled for each document. The word-level variables *z*_*dn*_, *w*_*dn*_, are sampled in each text-document for each word.

LDA portrays documents as a mixture of topics and topics as a mixture of words. It works by going through each document (d) and randomly assigns each word (w) in a document to one of the k topics (chosen beforehand).

The general working of the algorithm can be explained with the help of the following equation: (2)

*where t represents a specific topic

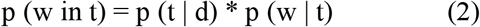

where we can define the following terms:

- p (t | d) represents the proportions of words in d documents assigned to the topic t
- p (w | t) represents the proportions of assignment s to topic t over every document, captures the documents in topic t because of word w.

If a word (w) has a high probability of being in a topic (t), all documents having the respective word will be associated with the topic. Similarly, suppose w is not probable to be in “t”. In that case, the documents which contain w will have a low probability of being in “t” because the rest of the words in the document will belong to another topic. Hence, the document will have a higher probability for those topics.

### 3.3. Sentiment Analysis and Comment Mining

Sentiment analysis is the classification of emotions within data using text analysis tools that automatically mine unstructured data for opinions and emotions, harnessing the power of deep learning to understand text beyond simple definitions. First, an appropriate lexicon was used for rule-based sentiment analysis, specifically attuned to sentiments expressed in social media. I chose the VADER lexicon[14], empirically validated by judges; VADER incorporates a “gold-standard” sentiment lexicon that is especially attuned to microblog-like contexts. VADER scores a sentence by comparing key words against its dictionary and its text pre-processing, further refined by making tweaks to the source code to handle exceptions.

#### Algorithm 2: Topic Recommendation

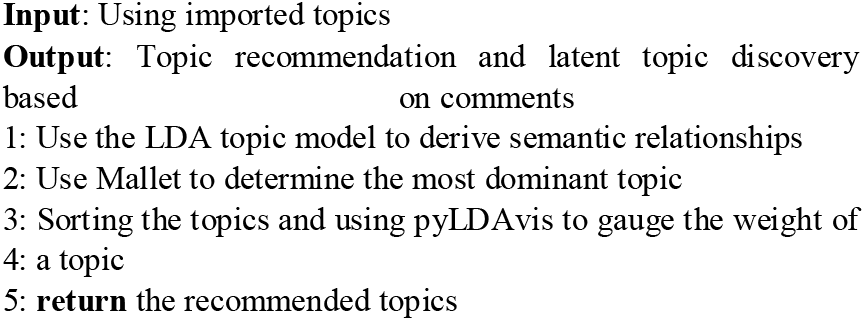

The use of Gibbs sampling makes the time complexity for the LDA operation of the order O(N K), N being the size of the corpus (Reddit comments), and K representing the topic number. The first four lines of the algorithm show the process of extracting latent topics and suitably arranging data to help build semantic relationships.

The comments and headlines were analysed to understand public sentiments with visualisation tools and algorithms discussed in section 4.

### 3.4. Deep-Learning and Sentiment Classification

Deep learning[15], often used interchangeably with machine learning, is an evolution of the latter which chains together algorithms that simulate the human brain, otherwise known as artificial neural networks, to solve complex problems and tackle massive amounts of data. The main advantage of using artificial neural networks is their ability to model high-level abstractions and decrease dimensionality by using multiple processed layers based on complex structures or non-linear transformations. RNNs, a class of artificial neural networks and a cornerstone in most NLP works [16] – [18] offer an innate ability to use consecutive information and store previous calculations by augmenting a memory’s output function that uses previous outputs and saves calculations. Basic RNNs cannot learn long-term dependencies due to gradient vanishing or exploding, which transformer-based models have overcome, hence changing the landscape of NLP. Models such as BERT, RoBERTa[19] and ALBERT[20] are based on the same principle – training bidirectional transformer models on massive unlabelled text corpora, done using mask language modelling (MLM), next sentence prediction (NSP) and sentence order prediction (SOP). Models differ in how these methods are applied, but in general, training is done in an unsupervised manner.

redBERT is based on the BERT-LARGE (English, uncased, whole word masking) model. BERT-LARGE is trained mainly on raw text data from Wikipedia (3.5B words) and a free book corpus (0.8B words) [2]. To improve performance in subdomains, we have seen numerous transformer-based models trained on specialised corpora. BIOBERT [21] and SCIBERT [22] are trained using the same unsupervised training techniques as the main models (MLM/NSP/SOP). They can be trained from scratch, but it requires a substantial corpus, so a more common approach is to start with the trained weights from a general model. This process is called domain-specific pretraining, and when trained, such models can be used as replacements for general language models and be trained for downstream tasks. The processed text was converted to fixed dimension vectors by pre-trained embeddings, and the last dense layer passed through a softmax function for a single classifier output. BERT’s mechanism revolves around implementing a separate attention mechanism[23] on each layer of the model to let each token from the input sequence focus on any other token, described by:

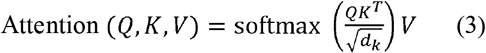

Where the keys(k), queries(Q), and values(V) are feed-forward neural network layers.

At the end of the framework, redBERT, a transformer-based model based on BERT, was used to assess the COVID-19 related comments of online users posted on Reddit to recognise the emotions/sentiment elicited from these comments.

## 4. Experiment Details

This section provides an in-depth account of the research process.

### 4.1. Classification

The first step of scrapping Reddit began with PRAW[24], a python package that lets us access Reddit’s API, enabling communication with Reddit and opening the door to access various subreddits and posts made across the platform. After registering, the subreddits to be analysed were chosen. After cleaning words from the headlines, as shown in algorithm-1 SIA[25], a sentiment analyser was used to determine the headlines’ polarity by assigning a negative, neutral, and positive value. The polarity of words was determined with the help of the VADER lexicon, which associates a sentiment rating to each word followed by TextBlob’s[26] use of a machine learning approach (Naive Bayes) to ‘teach’ the computer what words are associated with a negative or positive rating. While VADER focuses on content found everywhere, TextBlob benefits from being trained on a specific domain, enhancing our model’s polarity score.

The headlines’ compound polarity score can be easily calculated with an assumption that the values between 0.2 and −0.2 are considered neutral. The higher the margin, the larger the number of neutral headlines. The unavailability of a custom lexicon or manually labelling the data was overcome with the help of VADER and TextBlob.

After classifying the headlines using various python techniques, word clouds(Fig.3) and word frequency distribution curves were used to study patterns in the data and visualise the gradual change in data throughout this study.

### 4.2. Topic Discovery

Searching for latent topics and developing relationships was done with the help of LDA and Gibbs Sampling. After tweaking, the number of topics and words was set to 5 and 10, respectively. The words in each topic were tokenised with the help of Gensim’s simple_preprocess() and grouped into bigrams and trigrams[27], which are 2 and 3 frequently occurring words, respectively, using a Phrases model. This is followed by mapping each word with Gensim’s id2word and further interpreting keywords as an interactive web-based visualisation with pyLDAvis[28].

Bigrams and Trigrams, a subclass of N-gram models, help give context to words. These models predict the occurrence of words based on the occurrence of its N-1 previous words. As the conditional probability of up to N-1 words is too complex to calculate, a simplification assumption is introduced called Markov[29] assumption (equation 4), which approximates the history of a word to the last context the word.

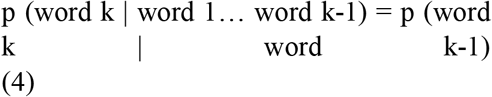

*where each word belongs to the same sentence.

After the data was ready to train, the LDA model was built after specifying the number of documents used in each training chunk and the total number of training passes. Fig.4 offers a look at a visualisation of the LDA model with the help of pyLDAvis.

The big bubbles scattered throughout the chart signify a topic. The coherence score of this topic model is 0.7140. The topic model was further improved by using MALLET’s[30] version of the LDA algorithm, which brought the coherence score to 0.73 by choosing an optimal number of topics, i.e., 20 in this case. Finally, each document’s topic distribution was discovered and used to infer the dominant topic in a document to gauge how widely a topic was discussed, the results of which are discussed in section 5.

### 4.3. redBERT

This model incorporates latent topic discovery and uses the relationship between different words to improve public sentiment polarity regarding COVID-19 comments in Reddit. redBERT builds on the transformer approach of forming relations between words with the help of the results obtained from topic modelling. To automatically classify the comments in the dataset, each comment was labelled as positive, negative, and neutral based on the sentiment score evaluated using the Sentistrength method. Based on Sentistrength, it was inferred that comment was negative if the negative sentiment was more significant than the positive sentiment score. The same was applied to determine the positive sentiment. For example, a score of +2 and −3 indicates negative polarity due to a net result of −1. Approximately 100000 comments were mined every month and broken down into a test/train ratio of 2/8. The proposed redBERT model was used along with fine-tuning the hyperparameters using NLP Architect by Intel AI Lab[31]. This model achieved an accuracy of 86.05%, which is higher than the traditional machine learning models.

## 5. Discussion and Analysis

Word clouds are a group of words depicted in different sizes where the size and boldness of words signify their importance within a given text. Fig.3 represents the word cloud of one of the topics discovered from the LDA process. The word cloud and sentiment analysis (the results shown in Fig.2) emphasise an upsurge of negative words than positive words.

Fig.4: The interdependency of different topics. Each circle represents a topic (the number of which is determined by the model’s coherence score), and the space between the circles signifies the broad range of topics discovered. Fig.5 offers a detailed analysis of the topic highlighted (topic 1) in Fig.4 and is used to determine the most dominant word in a given topic, helping understand the emotions conveyed by different users.

**Fig 2:**
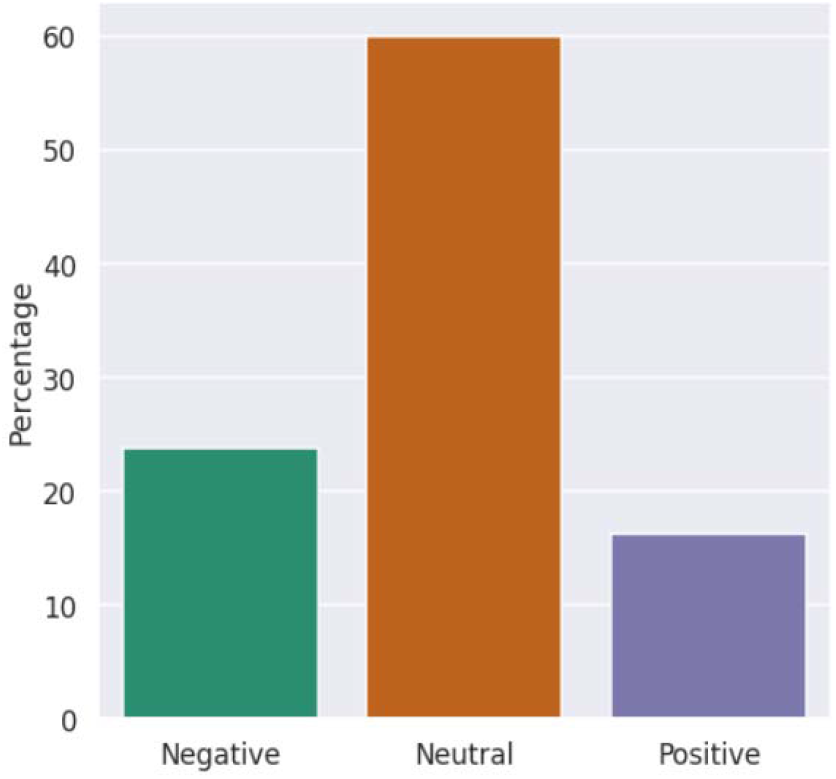
Sentiment graph of the percentage distribution of words.

**Fig 3:**
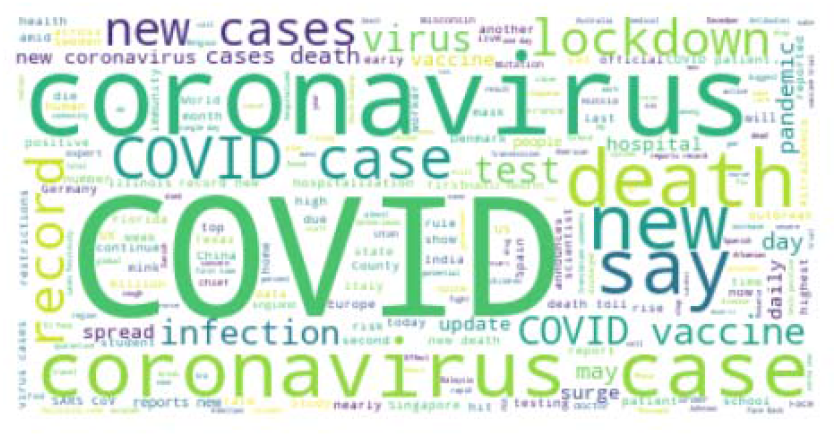
Word cloud visualisation based on word weights.

**Fig 4:**
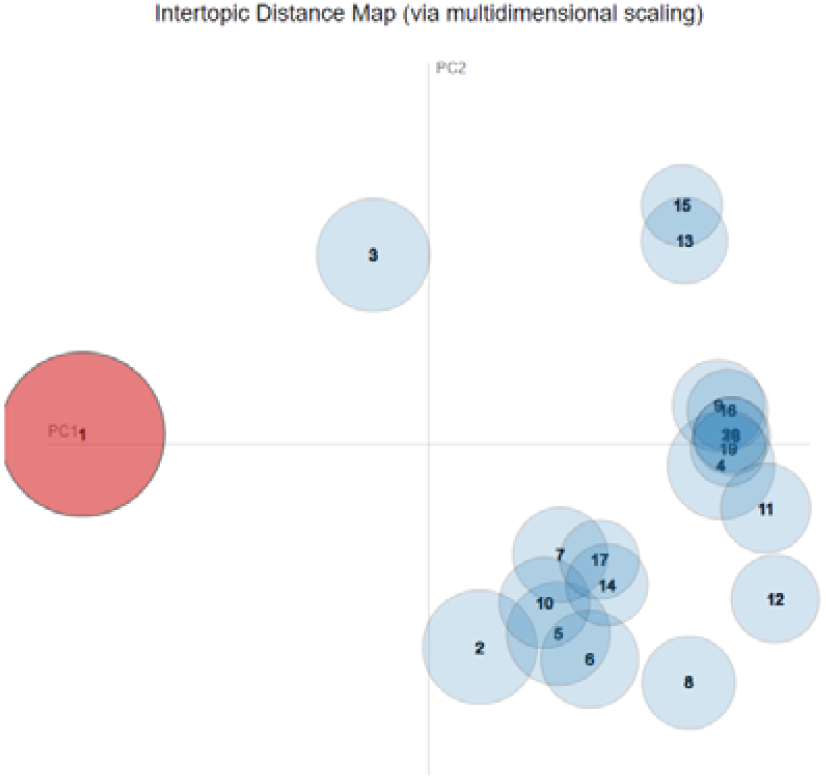
Topic distribution map of the topics discovered by LDA.

**Fig 5:**
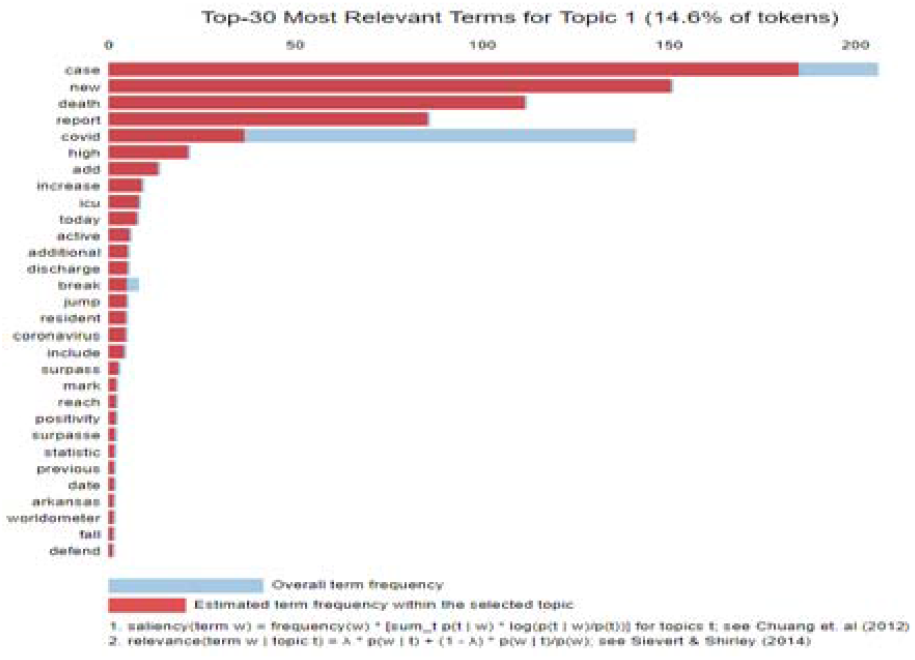
Distribution of the most dominant words in a topic.

The table in Fig.6 shows the most dominant words in a topic in the form of keywords that can build a general understanding of a topic. The percentage distribution of each word is shown in Fig.7.

For example, document no. 2 (Fig.6) is characterised by the words, “report”, “case”, “daily”, “theories”, “suggestion”, implying a growing awareness among people as they try to develop an understanding of the events transpiring around them. Each word’s percentage weightage is shown in Fig.7, which helps semantically identify what each piece of text is conveying.

**Fig 6:**
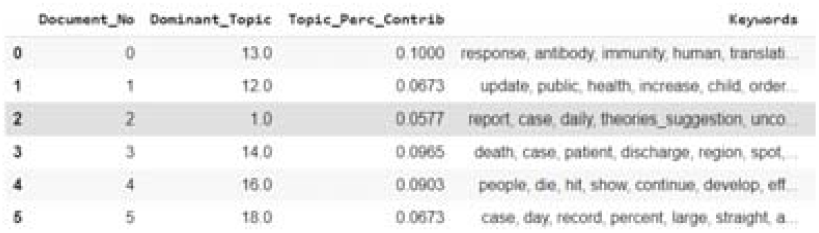
keywords and percentage contribution of dominant topics in a document.

**Fig 7:**
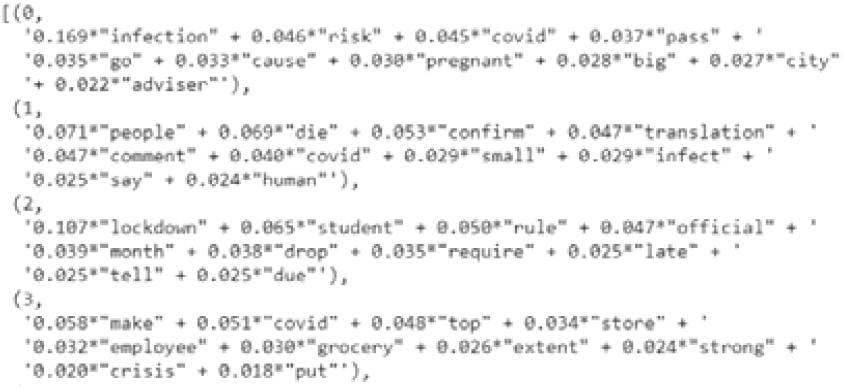
Percentage distribution of words in each topic.

Analysing social media platforms’ comments provides an unfiltered look into people’s outlook, which may be challenging to achieve through traditional techniques. The analysis was extended to check if a dependency could be found in the semantic aspects of user-comments of various issues on COVID-19 related topics. The dataset at the time of compilation included comments from 10 various subreddits. Meaningful latent topics about COVID-19 related issues were detected. A variety of different visualisations were used to interpret the generated LDA results. When LDA is applied to documents, it hypothesises a fixed set of topics, where each topic represents a set of words and maps the target documents under these imaginary topics. Integrating existing labelled knowledge sources improves this probabilistic model.

The top-ranked COVID-19 comments help recognise many words related to the needs and highlight discussions of the people or users on Reddit.

This research was limited to the English language; therefore, the results do not encompass sentiments in other languages. Moreover, this study was limited to the comments retrieved between January 20, 2020, and November 20, 2020; the gap between analysis and compilation may have affected the outcome. Overall, the study proposes that combining NLP and deep learning methods based on topic modelling and the BERT model enabled us to generate valuable information from COVID-19 related comments. Automated analysis of the mined comments can be used to determine online communities’ positive and negative actions and help researchers understand people’s behaviour in critical circumstances.

In the future, I plan on evaluating other social media, like Twitter, by adopting hybrid deep learning techniques for retrieving latent topics from public comments.

## 6. Conclusion

This paper serves as a facet to showcase a novel application of the BERT model to assimilate meaningful latent topics and sentiment comment classification on COVID-19, which can be extended to other issues of varying degrees. The results accumulated will help understand people’s needs concerning the pandemic and provide insight into the human interactions over the internet. Moreover, these findings may help improve practical strategies for public health services and interventions related to COVID-19.

## Data Availability

The data was extracted from Reddit using a scraping algorithm.

